# Orthogonal Contributions of Genetic, Clinical, and Social Determinants of Health Risk Burdens on Alzheimer’s Disease Pathophysiology

**DOI:** 10.64898/2026.07.07.26357509

**Authors:** Meri Okorie, Xiaqing Jiang, Paulina Tolosa-Tort, Rakshya U. Sharma, Alexandra L. Clark, Kristine Yaffe, Jennifer S. Yokoyama, Shea Andrews, Health and Aging Brain Study - Health Disparities

**Affiliations:** Department of Neurology and Weill Institute for Neurosciences, University of California, San Francisco, San Francisco, CA, USA; Fein Memory and Aging Center, University of California, San Francisco, San Francisco, CA, USA; Department of Radiology and Biomedical Imaging, University of California, San Francisco, San Francisco, CA, USA; Department of Psychiatry and Behavioral Sciences, University of California, San Francisco, San Francisco, CA, USA; Department of Epidemiology and Biostatistics, University of California, San Francisco, San Francisco, CA, USA; Department of Psychology, University of Texas at Austin, Austin, Texas, USA; San Francisco Veterans Affairs Health Care System, San Francisco, CA, USA; The Center for Population Brain Health, University of California, San Francisco, San Francisco, CA, USA

## Abstract

**Importance:** Alzheimer’s disease (AD) arises from complex interactions among genetic, clinical, and social determinants of health (SDoH) risk factors, yet their independent contributions to underlying AD pathophysiology remain elusive.

**Objective:** To quantify the effects of risk factors across amyloid (Aβ)/tau, neurodegeneration, and cognition.

**Design:** Cross-sectional analysis using structural equation modeling (SEM).

**Setting:** Health and Aging Brain Study–Health Disparities (HABS-HD), a community-based cohort study.

**Participants:** A total of 2,276 participants with demographics, genetic, clinical, and biomarker data from the baseline visit.

**Exposures:** *APOE* ε4 carrier status, AD polygenic risk score (AD-PRS), clinical risk score (CogDRisk), and a social determinants of health (SDoH) latent score derived using factor analysis.

**Main Outcomes and Measures:** Latent variables representing Aβ/tau pathology (plasma pTau_181_, plasma pTau_217_/Aβ_42_, amyloid PET positivity, and global standardized uptake value ratio), neurodegeneration (plasma neurofilament light, cortical thickness, hippocampal volume), and cognition (memory, executive, and language tests) were modeled and regressed on AD latent variables using SEM adjusted for age, sex, genetic principal components, and spoken language.

**Results:** The total analytic sample included 2,276 participants (mean age: 65.3 ± 8.7; non-Hispanic White: 43.0%, non-Hispanic Black: 16.2%, and Latinx/Hispanic adults: 40.8%). *APOE* ε4 was strongly associated with worse Aβ/tau latent variable (β=0.31; p<0.001), with smaller but significant associations with neurodegeneration (β=0.085; p<0.001) and cognition (β=0.083; p<0.001). Higher AD-PRS was modestly associated with worse Aβ/tau (β=0.075; p<0.01) but was not associated with neurodegeneration or cognition. A higher clinical risk score was significantly associated with worse neurodegeneration (β=0.16; p<0.001) but not with Aβ/tau or cognition. Adverse SDoH was associated with worse neurodegeneration (β=0.071; p<0.05) and strongly associated with worse cognition (β=0.22; p<0.001), with no associations with Aβ/tau.

**Conclusion and Relevance:** Genetic risks were primarily associated with Aβ and tau pathology, clinical risks with neurodegeneration, and SDoH risks with cognition, suggesting that risk factors exert differential effects on AD pathophysiology. Future studies investigating additional risk factors and their longitudinal associations with AD pathophysiological changes are warranted.

**Key Points:** *Questions:* Do Alzheimer’s disease (AD) risk factors differentially influence underlying AD pathophysiological processes, and do these associations vary across demographic subgroups?

*Findings:* Genetic and genomic risk burdens demonstrated the strongest associations with amyloid and tau pathology; clinical risk burden with neurodegeneration; and adverse social determinants of health with cognition.

*Meaning:* Because of the differential effects of risk factors on the underlying AD pathophysiology, a one-size-fits-all approach to AD risk prediction and prevention is insufficient. AD risk assessment should leverage multidomain frameworks incorporating genomic, clinical, and social determinants of health information to better inform disease development and progression.

## 1. BACKGROUND

Alzheimer’s disease and Alzheimer’s disease-related dementias (AD/ADRD) are the most common neurodegenerative diseases among older adults and arise from multifactorial etiologies^1^. The development of AD is characterized by amyloid (Aβ) deposition, tau accumulation, neurodegeneration, and ultimately cognitive decline^2^. The risk for AD-related cognitive impairment is shaped by the interplay of genetic, clinical, and social determinants of health (SDoH) factors acting across the life course^3,4^. The ε4 allele of apolipoprotein E (*APOE* ε4) is the strongest genetic risk factor for late-onset AD^5^, with genome-wide polygenic risk scores (PRS) further capturing the highly polygenic architecture of the disease^3,6^. In parallel, 14 modifiable risk factors have been identified that account for up to 45% of dementia cases^7^, enabling the development of cumulative clinical risk scores (CRS). SDoH represents a broad set of environmental exposures, encompassing the social, economic, and environmental conditions in which individuals are born, grow, live, work, and age^8^. Approximately 15% of dementia cases are estimated to be attributable to SDoH^9^, and growing evidence implicates disadvantaged SDoH as fundamental drivers of cognitive aging and dementia disparities^10,11^. These sources of risk factors together contribute to AD/ADRD pathogenesis; however, it remains unclear how these risk factors jointly influence disease processes.

While genetic, clinical, and SDoH risk factors have largely been identified through their associations with cognitive and clinical outcomes, less is known about how they relate to specific AD-related pathophysiological processes^10–13^. Advances in biomarkers of AD pathology have enabled the characterization of how risk factors influence the pathological processes underlying AD using the Aβ, tau, and neurodegeneration (A/T/N) framework^14–16^. Biomarker studies have overlooked social and life-course exposures and have typically focused on a single biological pathway, limiting the understanding of these relationships^17–21^. In addition, studies have focused on individual components of SDoH rather than on modeling its multidimensional and cumulative effects on cognition and pathology^22–24^. Although some work has linked lifestyle and SDoH factors to AD biomarkers, such investigations remain limited, and only a few studies have evaluated the effects of multimodal risk burdens on AD pathology within a unified model^25–28^. Consequently, the field lacks an understanding of the extent to which genetic, clinical, and SDoH factors affect the underlying AD/ADRD pathophysiology.

We therefore sought to quantify the contributions of genetic (*APOE* ε4, AD-PRS), clinical (CRS), and SDoH risk factors to AD/ADRD-related pathophysiology—including amyloid, tau, neurodegeneration, and cognition—within a unified model in the racially and ethnically diverse Health and Aging Brain Study–Health Disparities (HABS-HD) cohort. Elucidating these relationships may improve our understanding the mechanisms of AD pathogenesis by identifying pathways in which these risk factors influence underlying disease processes and inform precision prevention strategies.

## 2. METHODS

### 2.1 Study Populations

The HABS-HD comprises non-Hispanic White (NHW), non-Hispanic Black, and Hispanic participants recruited at the University of North Texas Health Science Center, Fort Worth, Texas^29,30^. The study is an ongoing longitudinal, community-based cohort that has collected information on demographics, clinical history, lifestyle, SDoH, genetic data, and AD endophenotypes, including plasma biomarkers, neuroimaging, and neuropsychological tests. Study eligibility was restricted to generally healthy individuals without any major psychological or medical conditions who agreed to undergo sample collection and neuroimaging assessments (N=3,116). Of these, 840 participants were excluded from the main analysis due to missingness of at least one exogenous variable, including *APOE* genotype, PRS, genetic principal components (PCs), or the SDoH latent score, resulting in an analytic sample of 2,276 participants. All analyses were conducted using baseline data included in the HABS-HD. Written informed consent was obtained from all HABS-HD participants or their legally authorized representatives before study participation.

### 2.2 Biomarker Harmonization and Standardization

Fasting blood samples were processed according to international pre-analytical guidelines ^30^, with all samples processed and stored at −80°C within 2 hours of collection. Plasma samples were assayed using single-molecule array (SIMOA; Quanterix) technology and were quantified using commercially available assay kits following standardized laboratory protocols with quality assurance and QC procedures^30^. Biomarker processing and harmonization procedures have been described previously^31,32^. In brief, plasma biomarkers (Aβ42, pTau181, pTau217, NfL) were log-transformed and standardized. Neuroimaging measures included cortical thickness and hippocampal volume derived from structural MRI, and global amyloid PET SUVR normalized to the cerebellum (positivity threshold=1.08)^30^. Neuropsychological tests measuring memory, language, and executive function domains were z-score normalized.

### 2.3 Genetics

*APOE* genotype was determined using TaqMan genotyping assays. Genome-wide SNP genotyping was performed using the Illumina Global Screening Array (GSA), with standard variant and sample-level quality controls applied as previously described^31^. The *APOE* ε4 allele carrier status was encoded as a binary variable of ε4+ carriers (ε2/ε4, ε3/ε4, or ε4/ε4) and non-ε4 carriers. A cross-ancestry PRS was generated using PRS-CSx based on prior benchmarking of AD-PRS methods^31^. In brief, the auto-shrinkage parameter and meta-analysis option were enabled for PRS construction. Variants within the *APOE* region (±500 kb; GRCh37, chr19:bp44909053–45912650) were excluded from the scorefile. Individual-level PRS was generated for each participant using pgsc_calc^33^, with automatic liftover to GRCh38 and normalization by genetic ancestry.

### 2.4 Clinical Risk Burden

Clinical risk burden was assessed using Cognitive Health and Dementia Risk Reduction (CogDRisk)^34^, as previously described and benchmarked as the best-performing CRS model^32^. The CogDRisk score includes age stratified by sex, education, midlife (≤65 yrs) obesity, dyslipidemia, diabetes stratified by sex, history of stroke, history of TBI, hypertension, atrial fibrillation, clinical diagnosis of insomnia, depression, physical inactivity, cognitive engagement, social engagement, diet, and current smoking. Age, sex, and education were modeled separately as covariates. The final score was calculated for each participant and was standardized.

### 2.5 Social Determinants of Health

We utilized factor analysis to construct an SDoH latent variable and estimate participant-level latent scores, as described in the Supplemental Contents. The dataset was randomly split into non-overlapping 50/50 samples for exploratory factor analysis (EFA) and confirmatory factor analysis (CFA), with sampling stratified by age, sex, and self-reported race to ensure demographic balance. SDoH indicators included measures of social support, chronic stress, anxiety, socioeconomic status, healthcare access, and demographic characteristics. EFA was used to identify the latent factor structure, and the resulting three-factor model was confirmed using CFA with a second-order SDoH latent construct. Individual SDoH factor scores were extracted for downstream analyses.

### 2.6 AD Latent Variables

CFA was used to define three AD latent variables: Aβ/tau, neurodegeneration, and cognition. Aβ/tau was indicated by plasma pTau_181_, pTau_217_/Aβ_42_, Aβ PET positivity, and Aβ PET global SUVR. Neurodegeneration was indicated by plasma NfL, cortical thickness, and hippocampal volume. Cognition was indicated by eight neuropsychological tests. Prior to CFA, Aβ PET measures were residualized by regressing them on PET scanner type, and plasma biomarkers were residualized on BMI and eGFR.

### 2.7 Statistical Analysis

Structural equation modeling (SEM) was used to derive the AD latent pathology and to perform regression analyses to assess the effects of *APOE* ε4, AD-PRS, CogDRisk, and adverse SDoH latent score. AD-PRS, CogDRisk, and SDoH latent scores were all treated as numeric variables. SEM was estimated using maximum likelihood with full-information maximum likelihood (FIML) to handle missing data and adjusted for age, sex, spoken language, and genetic principal components (4 PCs). Model fit was assessed using the Comparative Fit Index (CFI), Tucker-Lewis Index (TLI), Root Mean Square Error of Approximation (RMSEA), and Standardized Root Mean Square Residual (SRMR) for all factor analyses. Thresholds of CFI and TLI ≥ 0.95 and RMSEA and SRMR ≤ 0.06 were used to indicate an acceptable/excellent model fit^35,36^.

### 2.8 Multigroup Analysis

To evaluate whether the associations between risk factors and the latent outcomes differed by sex and race/ethnicity, we conducted multigroup SEM. Separate multigroup analyses were performed for sex (male, female) and race/ethnicity (NHW, Black, and Latinx/Hispanic). For each grouping variable, we first estimated a multigroup SEM in which factor loadings were constrained to equality across groups while structural regression coefficients were freely estimated. This model was compared with a model in which both the factor loadings and all structural regression coefficients were constrained to equality across groups. Model comparisons were performed using likelihood ratio tests (LRT).

For racial/ethnic groups, a significant omnibus χ^2^ difference test was followed by a series of model comparisons to identify the source of heterogeneity. First, the structural paths corresponding to each predictor (AD-PRS, *APOE* ε4 status, CogDRisk, and SDoH) were freed across racial/ethnic groups, while all remaining regression coefficients were constrained to equality. LRTs were used to determine whether allowing the effects of an individual predictor to vary across racial groups improved model fit. For predictors showing racial heterogeneity, additional models were fitted to identify the latent outcome(s) contributing to group differences. For predictor–outcome associations demonstrating significant racial/ethnic heterogeneity, pairwise multigroup comparisons were performed by comparing nested models in which the predictor–outcome regression coefficient was constrained to be equal or freely estimated between the two groups, and the difference was tested using LRT. Pairwise p-values were adjusted for multiple comparisons using the Holm method. All model fits were evaluated using the χ^2^ statistic, TLI, CFI, RMSEA, and SRMR. Missing data were handled using FIML.

### 2.9 Sensitivity Analysis

FIML estimator in lavaan can be applied to missingness in exogenous variables in the regression models (e.g., age, sex, education). We performed full SEM using FIML for the exogenous variable for both the latent variables and the regression to assess the stability of FIML when including all participants (N=3,116).

All analyses were conducted using R version 4.5.3, with the lavaan package version 0.6.21.

## 3. RESULTS

### 3.1 Sample characteristics

A total of 2,276 participants from the HABS-HD cohort were included in the main analysis. The mean age was 65.4 (±8.7) years; 60.9% were female, and the mean years of education were 13.2 (±4.5) (Table 1, eTable 1). 416 participants were diagnosed with MCI, and 119 participants with dementia, with the prevalence of *APOE* ε4 carriers being the highest among participants with dementia (Table 1). A total of 3,116 participants were included in the sensitivity analysis. The mean age was 65.2 (±8.7) years; 62.2% were female, and the mean years of education were 13.3 (±4.3). In both groups, NHW participants were the majority (main: 43.4%; sensitivity: 38.3%), followed by Latinx/Hispanic (main: 41.0%; sensitivity: 37.3%) and Black participants (main: 15.6%; sensitivity: 24.4%) (eTable 1).

**Figure 1.**
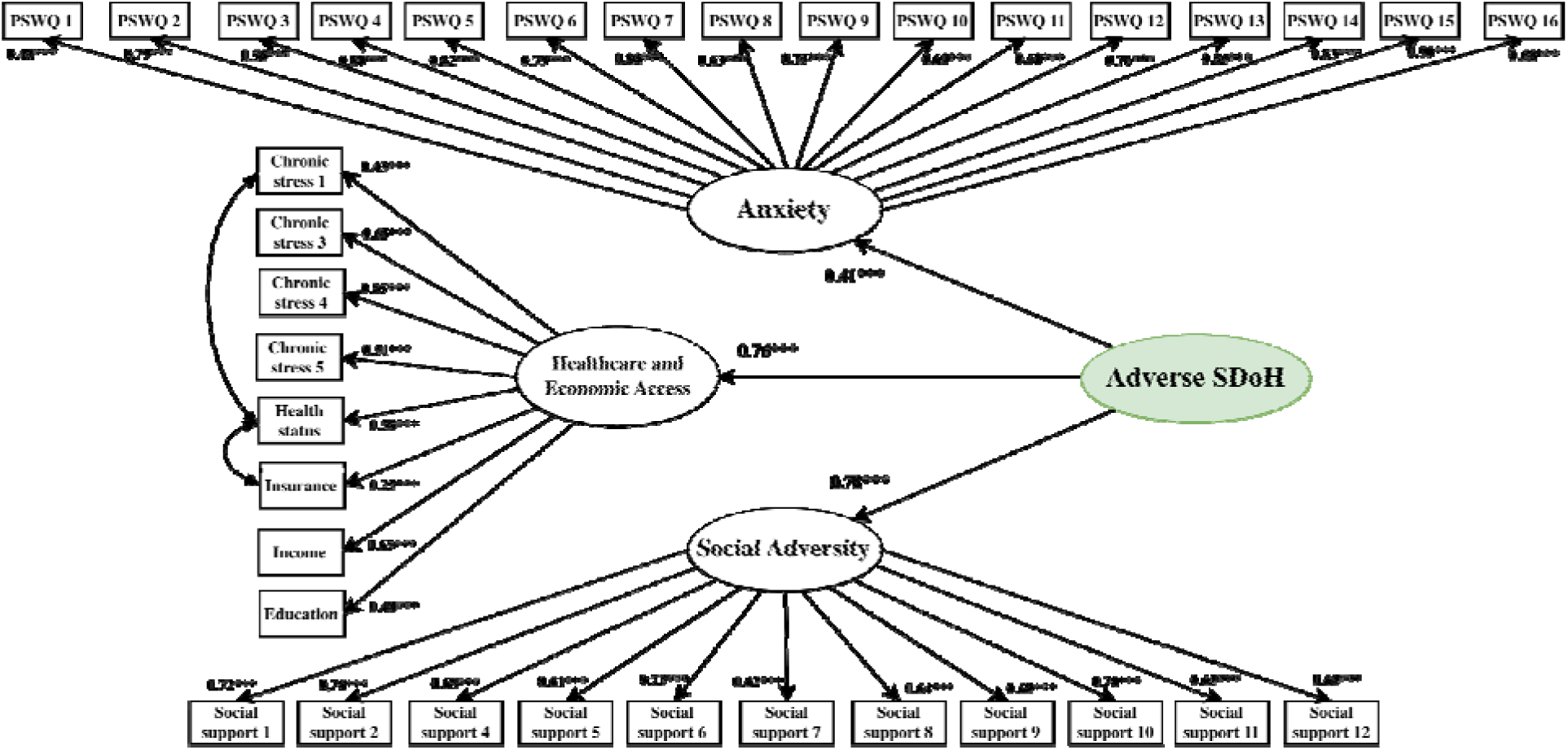
Latent structure of social determinants of health (SDoH). Second-order adverse SDoH latent structure was derived from exploratory and confirmatory factor analyses in the full cohort. The weighted least squared mean and variance adjusted (WLSMV) estimator was used for parameter estimation. The adverse SDoH latent score was estimated for 2,826 participants. Scaled CFI = 0.94, scaled TLI = 0.94, scaled RMSEA = 0.062 (90% CI: 0.060-0.063), scaled SRMR = 0.66. The b values indicate standardized path coefficients of indicators to corresponding latent variables. All displayed factor loadings were statistically significant. P-value thresholds: * < 0.05, ** < 0.01, *** < 0.001.

**Figure 2.**
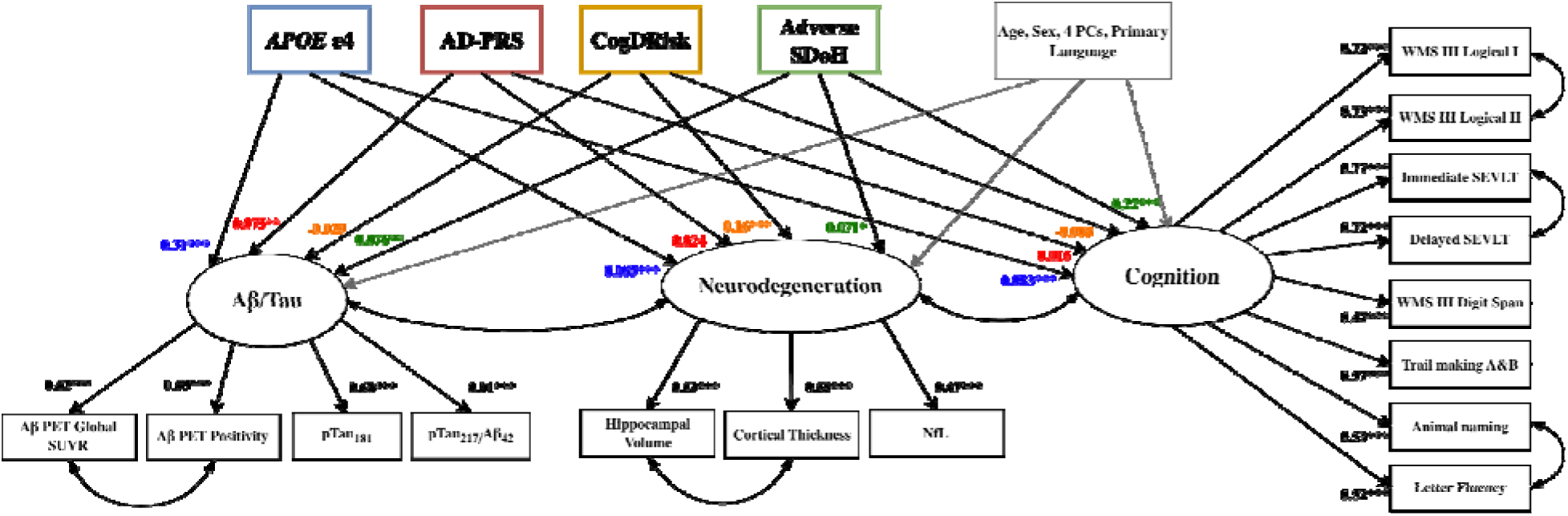
Structural equation modeling (SEM) examining the relationship between risk factors and Alzheimer’s disease (AD) pathology. The final SEM integrates all risk factors and AD latent variables. *APOE* ε4, AD polygenic risk score (AD-PRS), CogDRisk, and the adverse social determinants of health (SDoH) latent score were modeled as exogenous predictors and regressed on AD latent variables (Aβ/Tau, neurodegeneration, and cognition). All regression paths were adjusted for age, sex, four genetic principal components (PC1–PC4), and primary language. Model fit indices were: robust CFI = 0.92, robust TLI = 0.90, robust RMSEA = 0.054 (90% CI: 0.050–0.057), and SRMR = 0.044. Black β values represent standardized factor loadings of indicators onto latent variables, and colored β values represent standardized structural path coefficients from predictors to latent variables. All displayed factor loadings were statistically significant. Significance thresholds: *P < 0.05, **P < 0.01, ***P < 0.001.

**Table 1.**
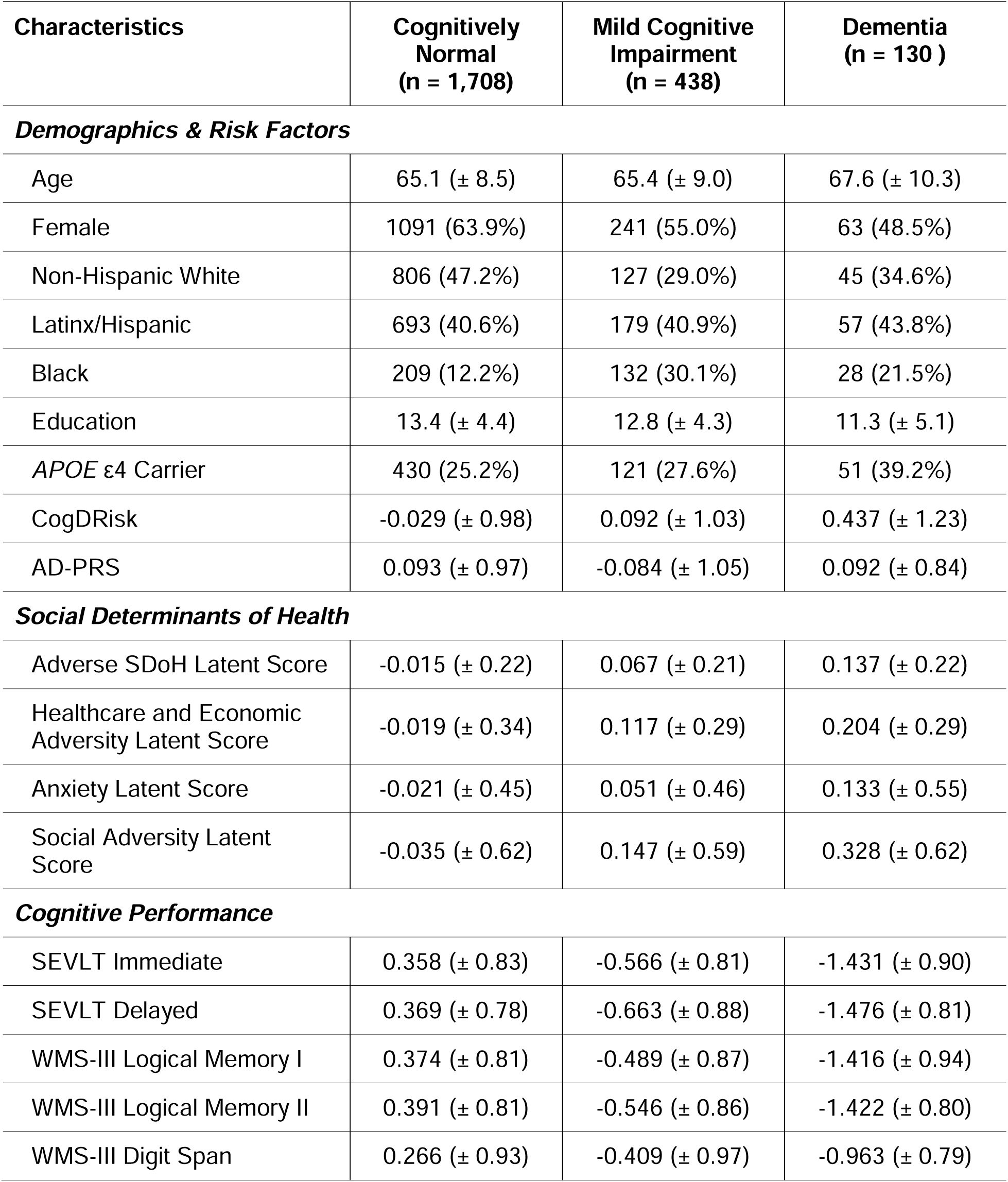
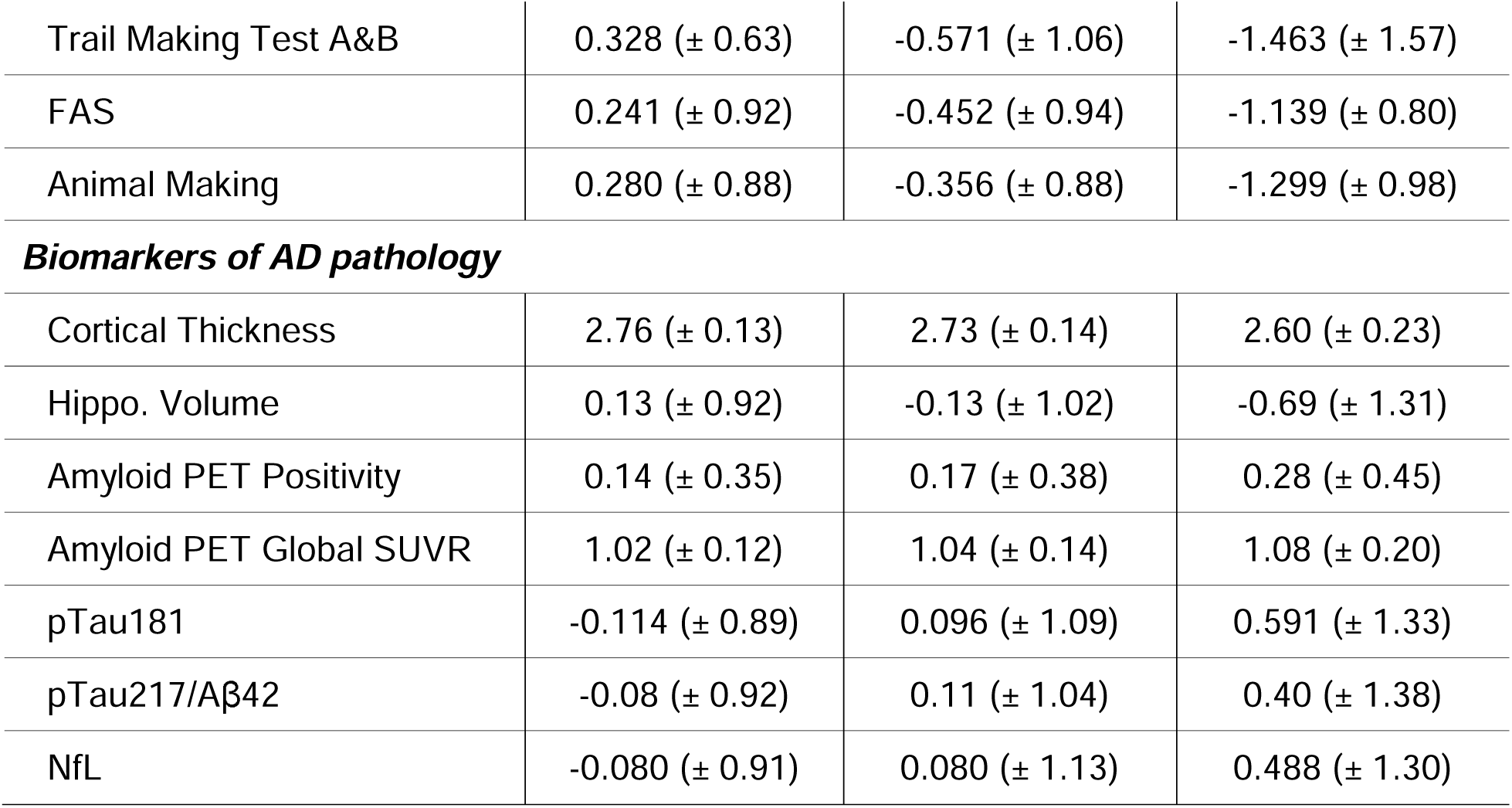
Descriptive statistics of the HABS-HD cohort used in the main analysis, stratified by cognitive status.

| Characteristics | Cognitively Normal<br>(n = 1,708) | Mild Cognitive Impairment<br>(n = 438) | Dementia<br>(n = 130) |
| --- | --- | --- | --- |
| **Demographics & Risk Factors** |  |  |  |
| Age | 65.1 (± 8.5) | 65.4 (± 9.0) | 67.6 (± 10.3) |
| Female | 1091 (63.9%) | 241 (55.0%) | 63 (48.5%) |
| Non-Hispanic White | 806 (47.2%) | 127 (29.0%) | 45 (34.6%) |
| Latinx/Hispanic | 693 (40.6%) | 179 (40.9%) | 57 (43.8%) |
| Black | 209 (12.2%) | 132 (30.1%) | 28 (21.5%) |
| Education | 13.4 (± 4.4) | 12.8 (± 4.3) | 11.3 (± 5.1) |
| APOE ε4 Carrier | 430 (25.2%) | 121 (27.6%) | 51 (39.2%) |
| CogDRisk | -0.029 (± 0.98) | 0.092 (± 1.03) | 0.437 (± 1.23) |
| AD-PRS | 0.093 (± 0.97) | -0.084 (± 1.05) | 0.092 (± 0.84) |
| **Social Determinants of Health** |  |  |  |
| Adverse SDoH Latent Score | -0.015 (± 0.22) | 0.067 (± 0.21) | 0.137 (± 0.22) |
| Healthcare and Economic Adversity Latent Score | -0.019 (± 0.34) | 0.117 (± 0.29) | 0.204 (± 0.29) |
| Anxiety Latent Score | -0.021 (± 0.45) | 0.051 (± 0.46) | 0.133 (± 0.55) |
| Social Adversity Latent Score | -0.035 (± 0.62) | 0.147 (± 0.59) | 0.328 (± 0.62) |
| **Cognitive Performance** |  |  |  |
| SEVLT Immediate | 0.358 (± 0.83) | -0.566 (± 0.81) | -1.431 (± 0.90) |
| SEVLT Delayed | 0.369 (± 0.78) | -0.663 (± 0.88) | -1.476 (± 0.81) |
| WMS-III Logical Memory I | 0.374 (± 0.81) | -0.489 (± 0.87) | -1.416 (± 0.94) |
| WMS-III Logical Memory II | 0.391 (± 0.81) | -0.546 (± 0.86) | -1.422 (± 0.80) |
| WMS-III Digit Span | 0.266 (± 0.93) | -0.409 (± 0.97) | -0.963 (± 0.79) |
| Trail Making Test A&B | 0.328 (± 0.63) | -0.571 (± 1.06) | -1.463 (± 1.57) |
| FAS | 0.241 (± 0.92) | -0.452 (± 0.94) | -1.139 (± 0.80) |
| Animal Making | 0.280 (± 0.88) | -0.356 (± 0.88) | -1.299 (± 0.98) |
| ***Biomarkers of AD pathology*** |  |  |  |
| Cortical Thickness | 2.76 (± 0.13) | 2.73 (± 0.14) | 2.60 (± 0.23) |
| Hippo. Volume | 0.13 (± 0.92) | -0.13 (± 1.02) | -0.69 (± 1.31) |
| Amyloid PET Positivity | 0.14 (± 0.35) | 0.17 (± 0.38) | 0.28 (± 0.45) |
| Amyloid PET Global SUVR | 1.02 (± 0.12) | 1.04 (± 0.14) | 1.08 (± 0.20) |
| pTau181 | -0.114 (± 0.89) | 0.096 (± 1.09) | 0.591 (± 1.33) |
| pTau217/Aβ42 | -0.08 (± 0.92) | 0.11 (± 1.04) | 0.40 (± 1.38) |
| NfL | -0.080 (± 0.91) | 0.080 (± 1.13) | 0.488 (± 1.30) |

### 3.2 Structural equation modeling reveals distinct contributions of risk factors across AD latent variables

The full SEM (N=2,276) was specified to examine the effect sizes of *APOE* ε4, AD-PRS, CogDRisk, and latent SDoH risk factors on AD latent pathology. The SEM included three endogenous latent outcomes: Aβ/tau, neurodegeneration, and cognition, with covariances specified between Aβ/tau and neurodegeneration, and between neurodegeneration and cognition. All the indicators loaded onto corresponding latent variables strongly and were statistically significant, with loadings ranging from 0.47 (WMS III Digit Span) to 0.91 (pTau_217_/Aβ_42_). The model demonstrated good global fit: robust CFI=0.92; robust TLI=0.90; robust RMSEA=0.054 (90% CI: 0.050–0.057); SRMR=0.044 (eTable 4). Local fit diagnostics revealed no substantial standardized residuals, and the retained model was selected based on parsimony, theoretical coherence, and model fit.

For the Aβ/tau latent variable, *APOE* ε4 had the highest effect on the latent variable (β=0.31, SE=0.035, p<0.001), whereas AD-PRS showed a modest association (β=0.075, SE=0.015, p=0.002). CogDrisk was not associated, whereas the adverse SDoH was (β=0.076, SE=0.069, p=0.002). For the neurodegeneration latent variable, CogDRisk was strongly associated with worse neurodegeneration (β=0.16, SE=0.012, p<0.001), along with *APOE* ε4 (β=0.085, SE=0.025, p=0.001). The adverse SDoH latent construct was also associated with worse neurodegeneration (β=0.071, SE=0.055, p=0.014), whereas AD-PRS was not. For the cognition latent variable, *APOE* ε4 was significantly associated, with an effect size similar to that observed with the neurodegenerative latent variable (β=0.083, SE=0.035, p<0.001). Adverse SDoH was strongly associated with worse cognition (β=0.22, SE=0.077, p<0.001), whereas AD-PRS and CogDRisk showed no associations with cognition (eTable 4).

Loadings of indicators onto corresponding latent variables remained consistent across AD latent variables in the sensitivity analysis (N=3,116) (eTable 5).

### 3.3 No sex differences in associations between risk factors and AD latent variables

To assess whether associations between the risk factors and AD latent variables differed by sex, we performed multigroup SEM comparing models with freely estimated versus constrained structural paths. Constraining the regression paths did not alter model fit (Δχ^2^(30)=35.80, *p*=0.22), indicating no sex differences in the associations of *APOE* ε4, AD-PRS, CogDRisk, or SDoH with Aβ/tau, neurodegeneration, or cognition (eTable 6, eTable 7).

### 3.4 Racial/ethnic differences driven by APOE ε4 effects on Aβ/tau

Multigroup SEM demonstrated racial/ethnic heterogeneity in the associations between risk factors and AD latent variables (Δχ^2^(60)=110.79, p<0.001). Sequential model comparisons showed that this heterogeneity was driven solely by *APOE* ε4 (Δχ^2^(6)=22.50, p=0.001), whereas no group differences were observed for AD-PRS, CogDRisk, or SDoH (eTable 7, eTable 8).

Further analyses showed that racial/ethnic heterogeneity was specific to the *APOE* ε4–Aβ/tau association (Δχ^2^(2)=20.46, p<0.001), with no differences for neurodegeneration or cognition (eTable 8). The *APOE* ε4 effect on Aβ/tau was strongest among NHW participants (β=0.37), followed by Latinx/Hispanic (β=0.27) and Black participants (β=0.21). Pairwise multigroup comparisons indicated that allowing the *APOE* ε4–Aβ/tau latent variable association to vary between NHW and Black participants improved model fit (Δχ^2^(1)=19.79, p_adj_<0.001), as did between NHW and Latinx/Hispanic participants (Δχ^2^(1)=9.09, p_adj_=0.005). In contrast, allowing the association to vary between Black and Latinx/Hispanic participants did not improve model fit (Δχ^2^(1)=1.99, p_adj_=0.16) (eTable 9).

## 4. DISCUSSION

In the community-based cohort of the HABS-HD study, we examined the relationships among genetic, clinical, and SDoH risks and the underlying AD pathophysiology, indicated by Aβ/tau, neurodegeneration, and cognition latent variables. *APOE* ε4 had the strongest effect on the Aβ/tau latent variable among other risk-pathology relationships. AD-PRS, on the other hand, also had a moderate effect on the Aβ/tau latent variable and was not significantly associated with neurodegeneration and cognition. The strong association of AD genetic risks with more upstream of the disease pathology recapitulates prior reports of the genetic impacts on AD pathology^37–39^. Given the cohort being generally healthy and cognitively unimpaired, it may have limited the enrichment of neurodegenerative markers and attenuated the associations of AD-PRS and neurodegeneration.

Many clinical risk scores, including CogDRisk, are derived using all-cause dementia as the outcome, which may explain their stronger association with neurodegeneration than with upstream Aβ/tau pathology or downstream cognition. This pattern is consistent with evidence that clinical risk burdens may contribute to cognitive impairment through neuronal injury and neuroinflammation (e.g., vascular comorbidities, metabolic dysfunction, and systemic inflammation) rather than through canonical AD pathology^40,41^. Similarly, the strong association between the cumulative SDoH score and cognition suggests that social determinants may influence cognitive reserve and performance independently of Aβ and tau pathology. Together, these findings indicate that different risk domains map onto distinct stages of the AD pathological cascade: genetic risk is most strongly associated with proteinopathy, whereas clinical and SDoH risk may act primarily through non-Aβ/tau neurodegenerative mechanisms, with adverse SDoH exerting the strongest influence on cognition.

Multigroup analyses revealed differential associations between *APOE* ε4 and Aβ/tau latent variables across racial/ethnic groups, which were not observed for neurodegeneration or cognition, suggesting that racial differences in *APOE* ε4 effects may primarily operate through amyloid and tau pathology rather than downstream neurodegenerative or cognitive processes. The observed pattern of *APOE* ε4 effects is consistent with previous studies reporting the strongest associations among NHW individuals, attenuated associations among Black individuals, and intermediate effects among Latinx/Hispanic admixed populations^42–46^. The concordance of our latent variable approach with these established observations supports the validity of the AD latent variable construct and suggests that *APOE* ε4 exerts race/ethnicity-specific effects on AD pathology.

The study includes a few limitations. First, all analyses were cross-sectional, which restricts our ability to evaluate temporal relationships among multidomain risk factors, AD proteinopathy, and cognitive decline. Second, the construction of both the CRS and the SDoH latent variables was constrained by available data. In particular, the SDoH measure did not include several relevant domains (e.g., neighborhood and built environment, education and healthcare quality, food access), which may have limited the specificity of the SDoH construct. Finally, we also did not examine other AD/ADRD-related pathologies, such as cerebrovascular disease, Lewy body disease, TDP-43 proteinopathy, or α-synuclein pathology. Future studies could extend this multidomain framework to evaluate the associations of these pathologies with risk factors.

Despite these limitations, this study has several strengths. Prior work has largely evaluated genetic, clinical, and SDoH risk factors in isolation. In contrast, we examined these domains simultaneously within a unified structural model, enabling direct comparison of their associations with underlying AD pathology. Furthermore, we utilized previously benchmarked and validated AD-PRS and CRS and derived the SDoH construct using rigorous exploratory and confirmatory factor analytic methods. Stratified analyses further extend these findings across demographic groups.

## 5. CONCLUSION

In a racially and ethnically diverse aging cohort, HABS-HD, genetic, clinical, and SDoH risk burdens exhibited distinct associations with AD-related pathology. These findings support evidence that genetic, clinical, and SDoH risk factors influence distinct stages of AD pathophysiology, advancing our understanding of the biological mechanisms underlying the complex disease development.

## Supporting information

Supplementary Content

## Data Availability

All data are available online at https://ida.loni.usc.edu/

## ACKNOWLEDGEMENTS

The authors would like to thank all the participants, staff, researcher teams, and partners of the Health & Aging Brain Study - Health Disparities (HABS-HD). The HABS-HD is supported by the National Institute on Aging of the National Institutes of Health under Award Numbers R01AG054073, R01AG058533, R01AG070862, P41EB015922, and U19AG078109. The content is solely the responsibility of the authors and does not necessarily represent the official views of the National Institutes of Health. We gratefully acknowledge the contributions of our study partners and their families, whose help and participation made this work possible.

