## Supplementary Content for "Orthogonal Contributions of Genetic, Clinical, and Social Determinants of Health Risk Burdens on Alzheimer’s Disease Pathophysiology"

**Supplemental Online Content**

eMethods

eResults

eFigure 1. Distributions of latent variables, stratified by cognitive diagnosis

eFigure 2. Distributions of latent variables, stratified by self-reported sex

eFigure 3. Distributions of latent variables, stratified by self-reported race/ethnicity

eReferences

**eMethods**

**Genetic Quality Control**

Participants were genotyped on the Illumina Global Screening Array (GSA), and genotype data underwent rigorous quality control (QC) using an in-house Snakemake Pipeline. Variant-level QC excluded SNPs with call rate <0.95 or violating Hardy–Weinberg equilibrium (p < 1 × 10^-6^). Sample-level QC removed participants with call rates below 0.95, discordant sex based on X chromosome heterozygosity, excessive or insufficient heterozygosity, or cryptic relatedness. Relatedness was evaluated across and within cohorts via identity-by-descent (IBD) using KING^1^, and individuals with IBD proportion <0.1875 were excluded, corresponding to less than halfway between second- and third-degree relatives. Non-genotyped SNPs were imputed using the TOPMed Imputation Server^2^, performing ancestry-specific imputation with the full TOPMed reference panel (Version R3) to allow accurate R^2^ estimation^3^, with Eagle and Minimac used for phasing and imputation. After imputation, poorly imputed variants (R^2^ < 0.3) or rare variants (MAF < 0.01) were removed. Ancestry groups were merged for joint analysis, after which variants and samples with call rates below 95% were removed. The final dataset included 3,116 participants (~22 million high-quality SNPs per individual), 1,194 are non-Hispanic White, 1,162 are Hispanic/Latinx, and 760 are Black participants.

**Biomarker Harmonization and Standardization**

To characterize the A/T/N and cognitive measures for AD endophenotypes, the HABS-HD study extensively phenotyped plasma biomarkers (e.g., Aβ_42_, pTau_181_, pTau_217_, and NfL), brain morphometry, Aβ PET, and neuropsychological tests. For downstream analyses, we applied the following harmonization and standardization for each AD endophenotype.

*Plasma biomarkers*: Fasting blood samples were collected, and the levels of plasma biomarkers were quantified using commercially available kits, Quanterix, for all the participants of the HABS-HD^4,5^. Aβ_42_, pTau_181_, pTau_217_, and NfL values were natural log-transformed and standardized to z-scores (mean = 0, SD = 1) across the study samples. *Brain morphometry:* Cortical thickness was calculated as the surface area-weighted mean of the right and left entorhinal cortex, fusiform gyrus, and inferior and middle temporal cortices. Hippocampal volumes were defined as the mean of the right and left hippocampal volumes derived from T1-weighted MRI scans. *Neuropsychological testing:* Composite scores for memory, language, and executive function were generated by averaging z-score-normalized results from the neuropsychological battery. The memory domain was assessed using immediate and delayed recall from the Wechsler Memory Scale (WMS-III) Logical Memory and the Spanish-English Verbal Learning Test (SEVLT). The language domain was assessed using Letter Fluency and Animal Naming tests. The executive function domain was assessed using the WMS-III Digit Span and the Trail Making Test, Parts A and B. In addition, the Mini-Mental State Examination (MMSE) and Clinical Dementia Rating (CDR) scale were administered to all participants as part of the neuropsychological assessment. *Amyloid PET:* For amyloid positron emission tomography (PET) scan variables, the global standardized uptake value ratio (SUVR) was calculated by normalizing tracer uptake to the whole cerebellum. Amyloid PET positivity was defined using a SUVR threshold of 1.08.

**Polygenic Risk Score**

Genome-Wide Association Study Base Dataset

We leveraged summary statistics from the largest publicly available ancestry-specific genome-wide association studies (GWAS) of Alzheimer’s disease and related dementias (ADRD) currently available. These included cohorts of European ancestry (Bellenguez et al.^6^: 39,106 cases, 46,828 proxy cases, 401,577 controls; FinnGen Release 6: 7,329 cases, 131,102 controls), African ancestry (Kunkle et al.^7^: 2,748 cases, 5,222 controls), East Asian ancestry (Shigemizu et al.^8^: 3,962 cases, 4,074 controls), and Caribbean Hispanic ancestry (Columbia University Study: 1,088 cases, 1,152 controls). There are no overlapping participants between the cohorts used in the base GWAS summary statistics and the HABS-HD cohort.

PRS Scorefile Generation

A cross-ancestry PRS was generated using PRS-CSx, which enhances trans-ancestry polygenic prediction by jointly modeling GWAS summary statistics from multiple populations^9^. Based on prior benchmarking of AD-PRS methods, PRS-CSx demonstrated the strongest predictive performance^10^ and was therefore selected for the present analysis. The global shrinkage parameter (phi) was specified as “auto,” enabling automatic shrinkage and estimation of SNP effect sizes from the input datasets. The “meta” option was set to true to aggregate SNP effects across all the GWAS summary statistics through inverse-variance–weighted meta-analysis. Ancestry-specific AD GWAS summary statistics were used as input to construct the scorefile. Prior to PRS computation, SNPs within the APOE locus and surrounding regions (±500 kb; GRCh37, chr19:bp44909053–45912650) were excluded from the final scorefile.

PRS Calculation and Ancestry Normalization

Individual-level PRS was generated for each participant using pgsc_calc^11^. During PRS calculation, liftover was applied automatically to convert the scorefile to the GRCh38 genome build. To reduce ancestry-related confounding arising from differences in PRS distributions and to facilitate more accurate comparisons across ancestrally diverse populations^12^, pgsc_calc was used to normalize PRS by ancestry in the HABS-HD cohorts using a principal components analysis (PCA)-based approach^13^. PCA was first performed on the 1000 Genomes (1KG) reference panel to derive principal components (PCs) capturing global genetic variation. Target individuals were then projected onto these PCs onto the PCA space. A Random Forest classifier trained on the reference panel PCA loadings (default: 10 PCs) was then used to assign each target sample to the most similar reference population^14^.

Following PC projection and ancestry assignment, PRS were normalized within ancestry groups. In the first step, PRS were regressed on PC loadings, and the resulting residuals were used to center the PRS distributions at zero across ancestries (znorm1). In the second step, a regression on the squared residuals was performed to adjust for variance, yielding standardized PRS (znorm2). Notably, phenotypic information, including case–control status, was not incorporated at any stage of the normalization process. This approach removes PRS variation attributable to population stratification driven by allele frequency and linkage disequilibrium differences, while preserving GWAS-derived SNP weights and relative risk rankings within ancestry groups. As a result, the normalization enables meaningful intra-ancestry comparisons while retaining disease-relevant genetic risk information within PRS.

**Clinical Risk Score**

CogDRisk^15^ was used to calculated accumulated clinical risk burden associated with AD. We have also previously benchmarked four CRS, and CogDRisk demonstrated the highest model performance compared to the other CRS^16^. The CogDRisk score includes age stratified by sex, education, midlife (≤65 yrs) obesity, dyslipidemia, diabetes stratified by sex, history of stroke, history of TBI, hypertension, atrial fibrillation, clinical diagnosis of insomnia, depression, physical inactivity, cognitive engagement, social engagement, diet, and current smoker. Atrial fibrillation, insomnia, cognitive engagement, and diet variables were excluded as this information was not available in the HABS-HD. Age, sex, and years of education were excluded from the scoring and were instead modeled as independent covariates in the SEM.

Participants who scored at least 1 SD below the mean on the social support questionnaire score were classified as ‘lonely,’ and those above this threshold were classified as ‘not lonely’ for the CogDrisk scoring. For physical activity measures, participants who answered ‘yes’ to item 6 or item 7 on the RAPA questionnaire were classified as physically active, reflecting an activity level of >150 minutes per week of moderate-to-vigorous activity. Baseline BMI at visit 1 was used in place of midlife obesity/BMI, which was not available in this cross-sectional study. Insomnia, atrial fibrillation, cognitive engagement, and diet measures were excluded from the scoring due to unavailability in the HABS-HD study.

**Adverse Social Determinants of Health Latent Score**

Exploratory Factor Analysis

EFA was performed to identify latent constructs underlying candidate measures, which included a battery of social support questionnaires, a battery of chronic stress questionnaires, a battery of the Penn State Worry Questionnaire (PSWQ), insurance information, income, years of education, marital status, health status, years lived in the US, and measures of medical visits. Variables with extreme missingness (>50%) were removed. Everyday Discrimination Scale (EDS) and Women’s Health questionnaires were removed due to high missingness. Sampling adequacy and suitability were assessed by the Kaiser-Meyer-Olkin test and Barlett’s test of sphericity. Factor retention was determined by assessing the eigenvalue-greater-than-one rule and scree plots. Orthogonal (varimax), oblique (promax), and no rotations were used to examine the correlated factor structures. Variables exhibiting weak loadings (<0.2), high cross-loadings, or strong multicollinearity were iteratively removed to improve model stability and interpretability. The final EFA solution retained a parsimonious set of indicators reflecting three underlying latent factors.

Confirmatory Factor Analysis

CFA was performed on the latent structure identified by EFA to formally confirm its structure, with the other 50% of the dataset. Based on the EFA results, a three-factor first-order measurement model was specified, which the latent factor majoritively comprised of PSWQ questionnaire was called Anxiety (AX), the latent factor majorly comprised of social support questionnaire was called Social Adversity (SA), and the latent factor comprised of subset of chronic stress questionnaire, self-rated health status, and medical and economic access was called Healthcare and Economic Adversity (HEA). A second-order latent factor (SDoH) was specified to capture the three first-order domains (AX, SA, and HEA), using weighted least squares mean and variance adjusted (WLSMV) as the estimator. This stepwise framework represents SDoH as a higher-level construct encompassing multiple domains, and the latent value for SDoH was estimated for each participant for the downstream analysis.

**Alzheimer’s Disease Latent Variables**

Confirmatory Factor Analysis

CFA was used to construct the AD pathology latent variables, namely Aβ/tau, neurodegeneration, and cognition pathology latent variables. Aβ/tau latent variable was indicated by plasma pTau_217_/Aβ_42_ ratio, Aβ PET positivity, and Aβ PET global SUVR. Neurodegeneration latent variable was indicated by plasma NfL level, cortical thickness, and hippocampal volume, with covariance between cortical thickness and hippocampal volume. Cognition latent variable was indicated by the eight neuropsychological test battery, with covariances among tests within each cognitive function domain (i.e., memory, executive function, language). Prior to CFA, observed indicators were residualized with respect to selected covariates to remove variance attributable to these factors. Aβ PET measures were adjusted for PET scanner type, and plasma pTau_217_ and Aβ_42_ were adjusted for body mass index and estimated glomerular filtration rate to account for the distribution and clearance of the plasma biomarkers. CFA was then conducted on the residualized indicators to evaluate the latent structure independent of covariate effects.

**eResults**

**Samples used for EFA and CFA**

The analytic sample (N = 3,116; non-Hispanic White: 1,194 (38.3%), non-Hispanic Black: 760 (24.4%), and Hispanic/Latinx adults: 1,162 (37.3%)) was stratified by sex and race and randomly partitioned for EFA and CFA (N = 1,558, respectively), with comparable demographic distributions across subsamples. Mean age was 65 (±8.75) in the EFA and 65 (±8.67) CFA subsamples. Same sample was used for CFA analysis of AD latent variables.

**Missingness**

For SDoH construction, the percentage of missingness across modeled variables ranged from 0.1% (self-reported health status) to 4.6% (income), with an overall missingness rate of 0.44%.

In the EFA and CFA models, estimation proceeded using complete-case analysis through WLSMV default handling of missing categorical data.

In the full SEM, the extent of missingness varied more substantially across variables, ranging from 0% to 56.2%, with the highest levels observed for amyloid PET global SUVR and amyloid PET positivity. Other variables with notable missingness included the pTau_217_/Aβ_42_ ratio (31.4%), cortical thickness (23.2%), *APOE* genotype (20.6%), and plasma NfL (11.6%). Little’s test for missing completely at random (MCAR) was statistically significant, indicating that the assumption of MCAR was not met.

To further characterize the missing data mechanism, we conducted logistic regression analyses to evaluate whether missingness was consistent with a missing at random (MAR) assumption. Demographic variables (age, sex, PRS, and years of education) were included as predictors, and a binary indicator of missingness (0 = missing, 1 = observed) was specified as the outcome for those variables with high missigness. Missingness in cortical thickness, *APOE* genotype, and amyloid PET measures was significantly associated with all demographic predictors. In contrast, none of the predictors were associated with missingness in either plasma biomarkers. The absence of associations between demographic variables and plasma biomarker missingness likely reflects measurement and processing variability inherent to the HABS-HD dataset. To mitigate these effects, we applied standardization and harmonization procedures across plasma biomarkers and restricted analyses to measurements obtained using the Quanterix platform. Despite these steps, residual variability cannot be excluded. Importantly, exclusion of plasma biomarkers from the final model was not pursued, as this could introduce biological bias given their established relevance to AD pathology and the lack of alternative measurements. This consideration represents a limitation of the present analysis. Missing data were handled using full-information maximum likelihood (FIML), which leverages all available observations without imputation and yields parameter estimates under MCAR/MAR assumptions. Prior simulation studies demonstrate that FIML maintains robust performance and outperforms alternative methods even under moderate-to-high levels of missingness (e.g., ~40%)^17^.

**EFA of SDoH Latent Construct**

EFA analysis of SDoH indicators demonstrated excellent sampling adequacy (MSA = 0.93) and a significant Barlett’s test of sphericity (χ^2^(df = 990) = 23,679.12, p < .001). Medicare insurance status, marital status, and two chronic stress questionnaires were excluded due to low MSA (<0.7). Scree plot inspection supported retention of a three-factor solution. Factor correlations from the EFA showed nontrivial interrelations: Factor1–Factor2 = −0.34, Factor1–Factor3 = 0.045, and Factor2–Factor3 = −0.38, supporting the choice of an oblique rotation given conceptually expected correlations among anxiety, social adversity, and healthcare/economic access. Indicators reflecting routine medical checkup recency, unmet need for physician care, personal primary care provider status, and private, Medicare, and Medicaid insurance coverage were removed due to low primary factor loadings (< 0.2). In addition, years lived in the United States and primary language were removed because of moderate cross-loadings across multiple factors, indicating poor discriminant structure. Inspection of the residual correlation matrix indicated adequate local fit, with no large residuals (max |residual| = 0.20).

**CFA of SDoH Latent Construct**

The three-factor structure was subsequently evaluated using CFA in the independent subsample. Three first-order latent variables (AX, HEA, SA) were specified, along with a second-order SDoH factor loading on each first-order construct using the weighted least squares mean and variance adjusted (WLSMV) estimator, as variables were majoritively categorical and non-normal. Residual covariances were specified between health status and chronic stress questionnaire item #1 (“Have you had a serious ongoing health problem?”), and between health status and insurance coverage (binary indicator of any coverage vs. uninsured), based on conceptual overlap. Model identification was established using the marker-variable method (one indicator loading fixed to 1.0 per latent variable).

The CFA demonstrated acceptable-to-good global fit: CFI = 0.980, TLI = 0.977, RMSEA = 0.057 (90% CI: 0.054–0.060), and SRMR = 0.065. Local fit was adequate; standardized residuals ranged from 0 to 0.21. All standardized factor loadings were statistically significant (p < 0.001), and no improper solutions (e.g., negative variances, standardized loadings > 1.0) were observed. Standardized AX loadings ranged roughly from 0.46–0.85 (PSWQ items), HEA loadings ranged from ~0.32–0.93 (chronic stress, income, insurance), and SA loadings ranged from ~0.64–0.74 (social support items). A second-order SDoH factor loaded on HEA, SA, and AX, with standardized first-order loadings of 0.66, 0.87, and 0.44, respectively.

The finalized second-order SDoH model was then re-estimated in the full analytic sample (N = 2,826). Model fit remained close to the thresholds of excellent fit: CFI = 0.98, TLI = 0.98, RMSEA = 0.062, 90% CI 0.061–0.064, SRMR = 0.067, and standardized loadings were consistent with the confirmatory subsample (second-order SDoH loadings: HEA = 0.74, SA = 0.76, AX = 0.43) (eTable 2). Final SDoH latent factor scores were predicted for each participant and were extracted for the downstream analysis.

**CFA of AD Pathology Latent Variables**

Three latent variables, Aβ/tau, neurodegeneration, and cognition, were indicated by corresponding endophenotypes, and were estimated using FIML. The Aβ/tau latent variable was indicated by plasma pTau_217_/Aβ_42_ level, Aβ PET positivity, and Aβ PET global SUVR, the neurodegeneration latent variable was indicated by plasma NfL level, cortical thickness, and hippocampal volume, and cognition latent variable was indicated by memory, executive function, and language neuropsychological test battery. The total of 3,116 participants were included, and the model demonstrated good global fit: robust CFI = 0.98; robust TLI = 0.98; robust RMSEA = 0.042 (90% CI: 0.037–0.048); SRMR = 0.030. All indicators loaded significantly onto their respective latent construct, with moderate to strong loadings (Aβ/tau: 0.74–0.83; neurodegeneration: 0.48–0.56; cognition: 0.55–0.65) (eTable 3).

eFigure 1. Distributions of latent variables, stratified by cognitive diagnosis. Kolmogorov-Smirnov tests confirmed statistically significant deviation of distributions of Aβ/Tau, neurodegeneration, cognition, and adverse SDoH latent scores between MCI vs cognitively normal and dementia vs cognitively normal group. Top left: Aβ/Tau latent score; top right: Neurodegeneration latent score; bottom left: Cognition latent score; bottom right: Adverse SDoH latent score. (N = cognitively normal: 1,638; MCI: 416; dementia: 119)

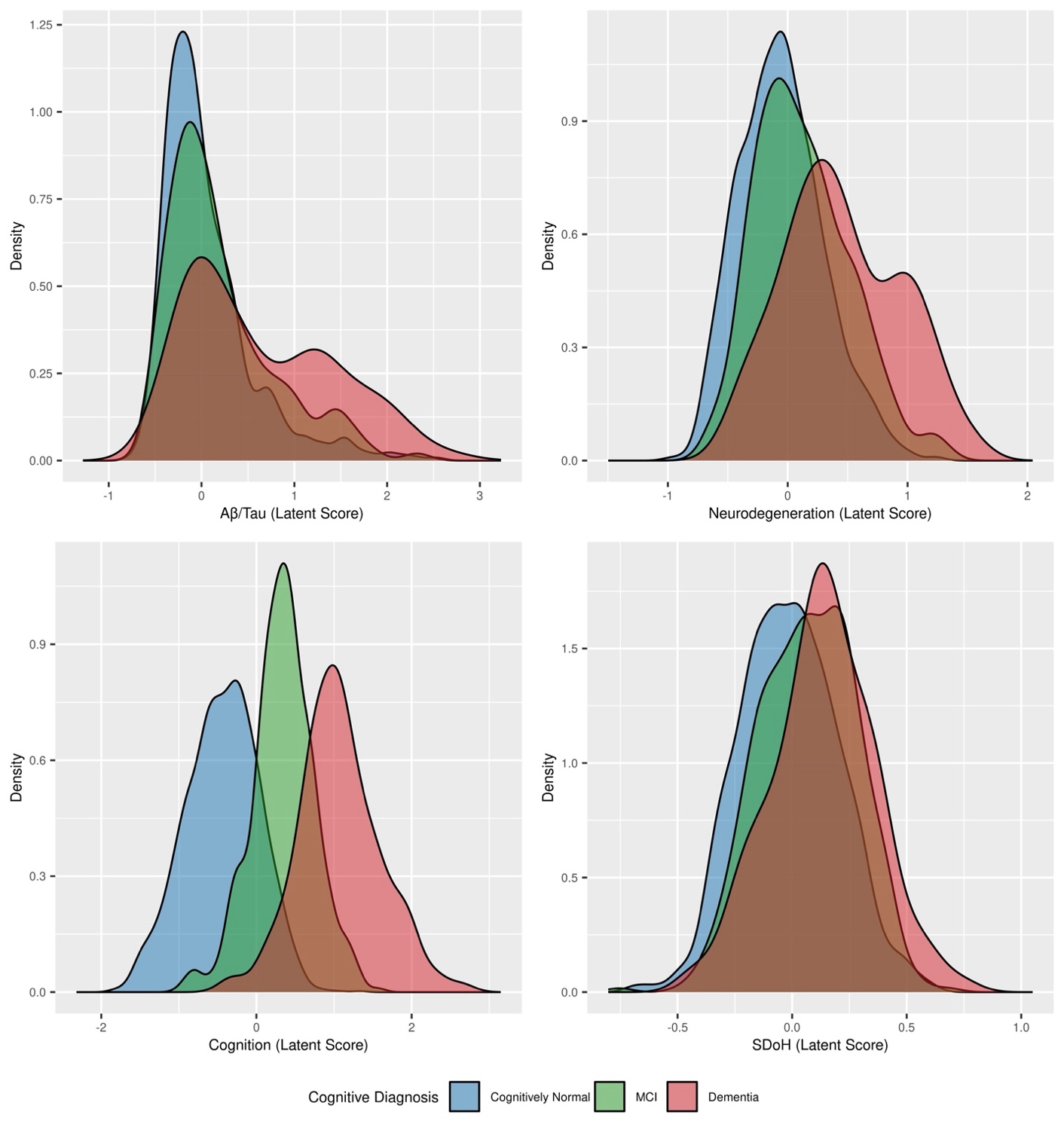

eFigure 2. Distributions of latent variables, stratified by self-reported sex. Kolmogorov–Smirnov tests indicated statistically significant differences in the distributions of Neurodegeneration and SDoH latent scores between sexes. Top left: Aβ/Tau latent score; top right: Neurodegeneration latent score; bottom left: Cognition latent score; bottom right: Adverse SDoH latent score. (N = Female: 1,323; Male: 850)

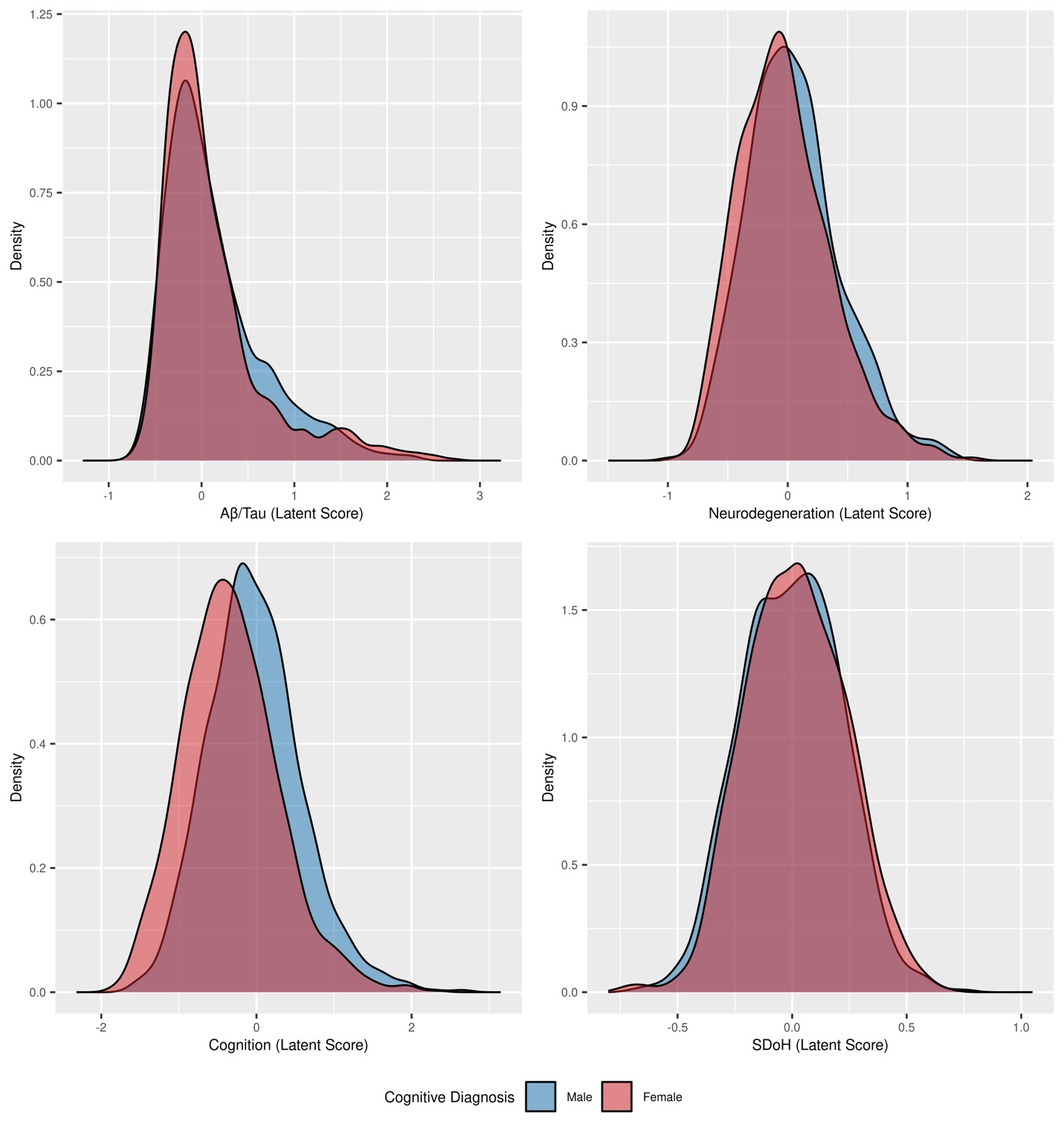

eFigure 3. Distributions of latent variables, stratified by cognitive diagnosis. Kolmogorov-Smirnov tests confirmed statistically significant deviation of distributions of Aβ/Tau, neurodegeneration, cognition, and adverse SdoH latent scores between NHW vs Black and Black vs Latinx/Hispanic participants. Top left: Aβ/Tau latent score; top right: Neurodegeneration latent score; bottom left: Cognition latent score; bottom right: Adverse SDoH latent score. (N = NHW: 944; Black: 338; Latinx/Hispanic: 891)

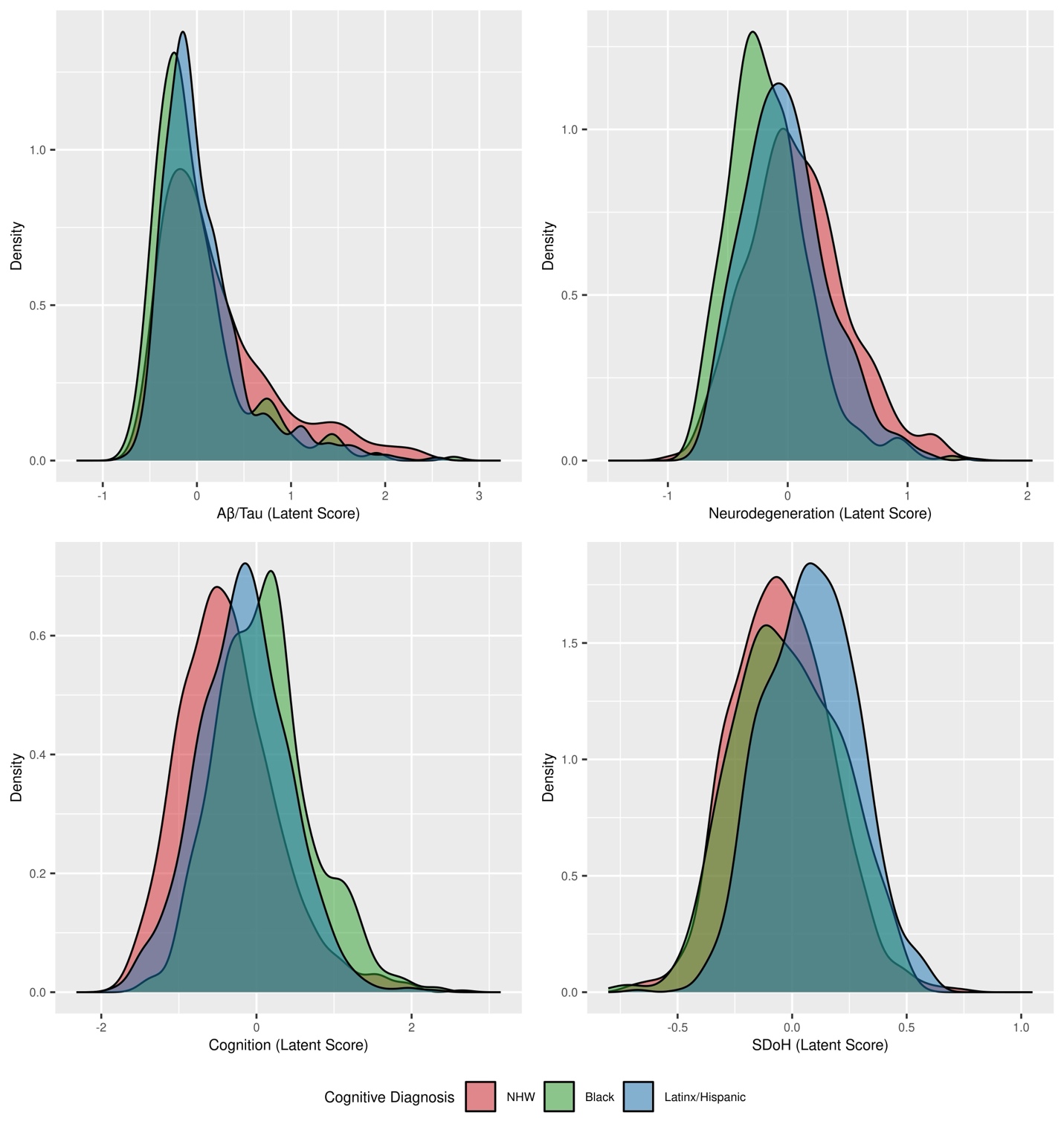

eReferences

1.	Relationship Inference in KING. https://www.kingrelatedness.com/manual.shtml.

2.	TOPMed Imputation Server. https://imputation.biodatacatalyst.nhlbi.nih.gov/#!
